# Prevalence and characteristics of medication errors at the outpatient settings, Jigme Dorji Wangchuck National Referral Hospital (JDWNRH): Retrospective study

**DOI:** 10.64898/2026.01.24.26344747

**Authors:** Kezang Tshering, Karma Rabgay, Kuenzang Rangdel Dorji, Khando Wangchuk, Thinley, Kharka B. Bhattarai, Ugyen Dema, Tshering Cheda, Tshering Dema Blon, Choeni Dema Dorji, Tashi Dema, Keran Neopany, Pelden Chejor

## Abstract

**Introduction:** Medication errors (MEs) are common events in the hospital that compromises the patient safety. Despite high prevalence of MEs being reported, there is a limited information at the outpatient settings.

**Method:** This study assessed the prevalence and characteristics of MEs by analyzing the data of pharmacist-led interventions recorded between 1^st^ November 2024 to 30^th^ September 2025 at the outpatient department of pharmacy, Jigme Dorji Wangchuck National Referral Hospital (JDWNRH).

**Results:** Of the 90108 prescriptions dispensed, 2045 (2.27%) MEs were recorded during the study period. Among which, 90.81%, 8.70% and 0.49% accounted to prescribing error, packaging error and dispensing error, respectively. More than half of the prescribing error were attributed with antibiotics (39.3%) and antihypertensives (13.9%) combined. Dosing error (63.5%) was highly prevalent type of prescribing error, followed by wrong frequency (12.0%), no indication (10.0%) and omission (5.7%). The severity of harm related to with prescribing error were observed as; no harm (212, 11.42%), minor (233, 12.55%), moderate (1364, 73.45%) and serious (48, 12.58%). Pharmacist-led interventions has prevented more than 90% of the potential harm related to the prescribing error.

**Conclusion:** Prescribing error is the most common type of MEs at the outpatient settings in JDWNRH. Majority of the prescribing errors were found to be moderately severe. Pharmacists-led interventions should be effectively integrated in the disease management system to enhance the overall patient safety. Concerned prescribers should enhance their knowledge, particularly in therapeutic management of infectious disease and hypertension to minimize the prescribing errors.

## Introduction

Medication errors (MEs) represent a common event in the hospital and pose a significant threat to patient safety. MEs occur during the medication management process involving prescribing, transcribing, administering and dispensing, which accounts for approximately one-quarter of all healthcare associated errors [1].

The World Health Organization (WHO) has estimated 1.3 million injuries annually and one death daily, associated with MEs in the United States (US) [2]. Hence, WHO has declared medication safety improvement as a third global patient safety challenge [3]. Following the WHO declaration, various interventions were proposed to mitigate MEs. Among which, a pharmacist-led intervention is one of the potential interventions recommended to combat MEs in the hospital settings [4].

Although MEs are extensively studied in inpatients care [5–7], there is limited evidence on the prevalence and nature of MEs in outpatient settings [8]. Despite limited information, four of every ten MEs were recorded in outpatient and ambulatory settings as reported by the National Health Services (NHS), England [9]. But MEs at the outpatient settings in the low-and middle-income countries remained poorly studied.

MEs are associated with significant healthcare cost. According to the National Patient Safety Agency, the estimated cost of preventable harm due to MEs was over *£*750 million annually in England [10]. This highlights the importance of understanding the prevalence and characteristics of MEs to adopt and implement targeted interventions to prevent MEs, particularly in the resource limited settings.

Adopting specific strategies based on the nature of MEs and its associated risk factors is recommended in preventing MEs [11]. Without understanding the prevalence and type of MEs at the specific healthcare settings, targeted interventions can’t be contextualized leading to higher risk of experiencing MEs. Therefore, our study aimed to understand the prevalence and characteristics of MEs at the outpatient setting by analyzing the data of pharmacist-led interventions maintained at the department of pharmacy, Jigme Dorji Wangchuck National Referral Hospital (JDWNRH), Bhutan.

## Method

### Design

A retrospective analytical study was conducted by analyzing the data of MEs recorded by the hospital Pharmacists between 1^st^ November and 30^th^ September 2025 at the Department of Pharmacy, JDWNRH.

### Study setting

Outpatient setting at JDWNRH consisted of clinical departments including Medical, Surgical, GOPD (General Outpatient Department), ENT (Ear, Nose, Throat), Orthopedic, Dermatology, Ophthalmology, Pediatric, Gynecology, Psychiatry, Dental, Forensic, Community Health Department and Emergency triage. The outpatient pharmacy at JDWNRH consisted of 10 dispensary counters with 9 hospital pharmacists and 28 Pharmacy Technicians (PT) involved in dispensing of prescriptions at the outpatient care.

### Data collection procedure

In August 2024, the department of pharmacy, JDWNRH has established a Pharmaceutical Care Intervention Unit (PCIU) consisting of one full-time hospital pharmacist to intervene on any outpatient prescriptions identified to contain any form of MEs.

All outpatient prescriptions generated through electronic Patient Information System (ePIS) were primarily reviewed for its appropriateness in terms of indications, dose, frequency and duration by the pharmacist. Omission, drug-drug interactions, de-prescribing and adverse drug reactions (ADRs) were also reviewed concurrently. In case of any prescribing error encountered, the concerned prescribers were contacted through telephone by the designated pharmacist at PICU for necessary amendments in the prescriptions. Following the interventions, description of MEs recorded includes pharmacist’s intervention note/recommendations, name of the medicine, type of MEs, prescriber category, department of the prescriber and the intervention outcome.

Packaging error in terms of wrong medicine being packed inside the dispensing envelope were identified and reported to PCIU by the dispensers. Additionally, dispensing error were recorded when the wrong medicines were dispensed and returned to the outpatient pharmacy by the patients. All MEs encountered and intervened are being recorded in the google sheet maintained at the PCIU.

While reviewing for appropriate indication, dose, frequency and duration of the treatment, prescribing compliance to both national and international evidence-based guidelines or references were considered by the hospital pharmacists. Some of the commonly referred international references included UpToDate, the British National Formulary (BNF), American Diabetes Association (ADA), KDIGO, NICE guidelines and Infectious Disease Society of America (IDSA). The available national references included National Antibiotic Guidelines-2018, National Essential Medicine Formulary (NEMF) – 2022 and Standard Treatment Guideline (STG) – 2014 of Bhutan. Medication reconciliation was performed to identify omission of medicines, no indication and the history of ADRs. Drug interaction checker in Medscape and Micromedex were used to check for the potential drug-drug interactions.

### Measures

The therapeutic categories were used to classify the individual medications. The overall prevalence of MEs included a sum of prescribing error, packaging error and dispensing error. In particular, the overall prescribing error includes if any one of the incidents such as no indication, omission, wrong indication, wrong dose, wrong frequency, wrong duration, wrong dosage form, no dose, no frequency, no duration, drug-drug interaction and ADRs were encountered.

The classification of potential harm associated with MEs was grouped as no harm, minor, moderate and serious based on the level of harm and patient care required. Classes of potential harm was defined as 1) No harm: MEs with no potential for patient harm, nor any change in patient monitoring; 2) Minor: MEs with potential for minor, non-life threatening, temporary harm that may or may not require any change in patient monitoring; 3) Moderate: MEs with potential for minor, non-threatening, temporary harm that requires patient monitoring; 4) Serious: MEs with potential for major, non-life threatening, temporary harm or minor permanent harm that requires high level of patient monitoring such as use of antidote. The classification of potential harm associated with MEs was adopted from Harm Associated with Medication Error Classification (HAMEC) tool developed by the expert panel consisting of medical clinicians, pharmacist and nurse [12]. In our study, MEs which have reached to the patients were considered as the actual harm.

### Data analysis

Data was validated, coded and exported to Statistical Package for the Social Sciences (SPSS) version 26 for analysis. Descriptive statistics such as frequency and percentage were used to describe the prevalence and characteristics of MEs.

### Ethical Approval

Considering no involvement of patient data, the study was exempted from the ethical approval by the Institutional Review Board (IRB), Khesar Gyalpo University of Medical Sciences of Bhutan (KGUMSB).

## Result

### Prevalence of ME

Of the 90108 prescriptions being dispensed, 2045 (2.27%) prescriptions were recorded with one type of MEs. The overall prescribing error was 2.06% (n=1857), followed by packaging error of 0.20% (n=178) and dispensing error of 0.01% (n=10) out of 90108 prescriptions dispensed.

From the overall ME, prescribing error, packaging error and dispensing error accounted to 90.81%, 8.70% and 0.49% respectively as shown in the **Figure 1**.

**Figure 1.**
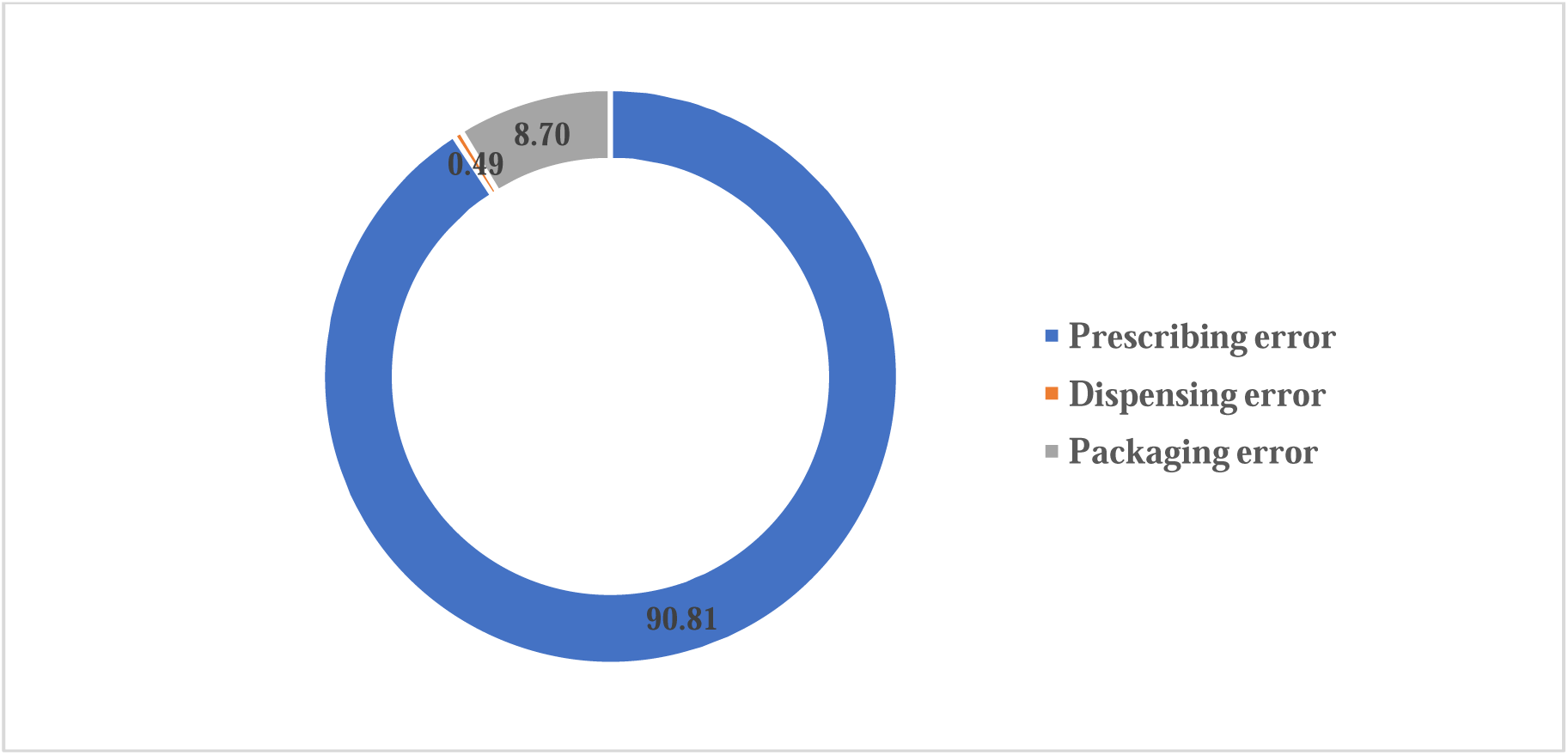
Proportion of medication errors at the outpatient setting, Jigme Dorji Wangchuck National Referral Hospital (JDWNRH) from 1^st^ November to 30^th^ September 2025.

### Characteristics of prescribing error

Based on the therapeutic classifications, more than half of the prescribing error recorded were attributed with antibiotics (39.3%) and antihypertensives (13.9%) combined as described in **Table 1**. The most common type of prescribing error was wrong dose (63.5%), followed by wrong frequency (12%), no indication (10.0%), omission (5.7%) and no dose (5.1%) as depicted in **Figure 2**.

**Figure 2.**
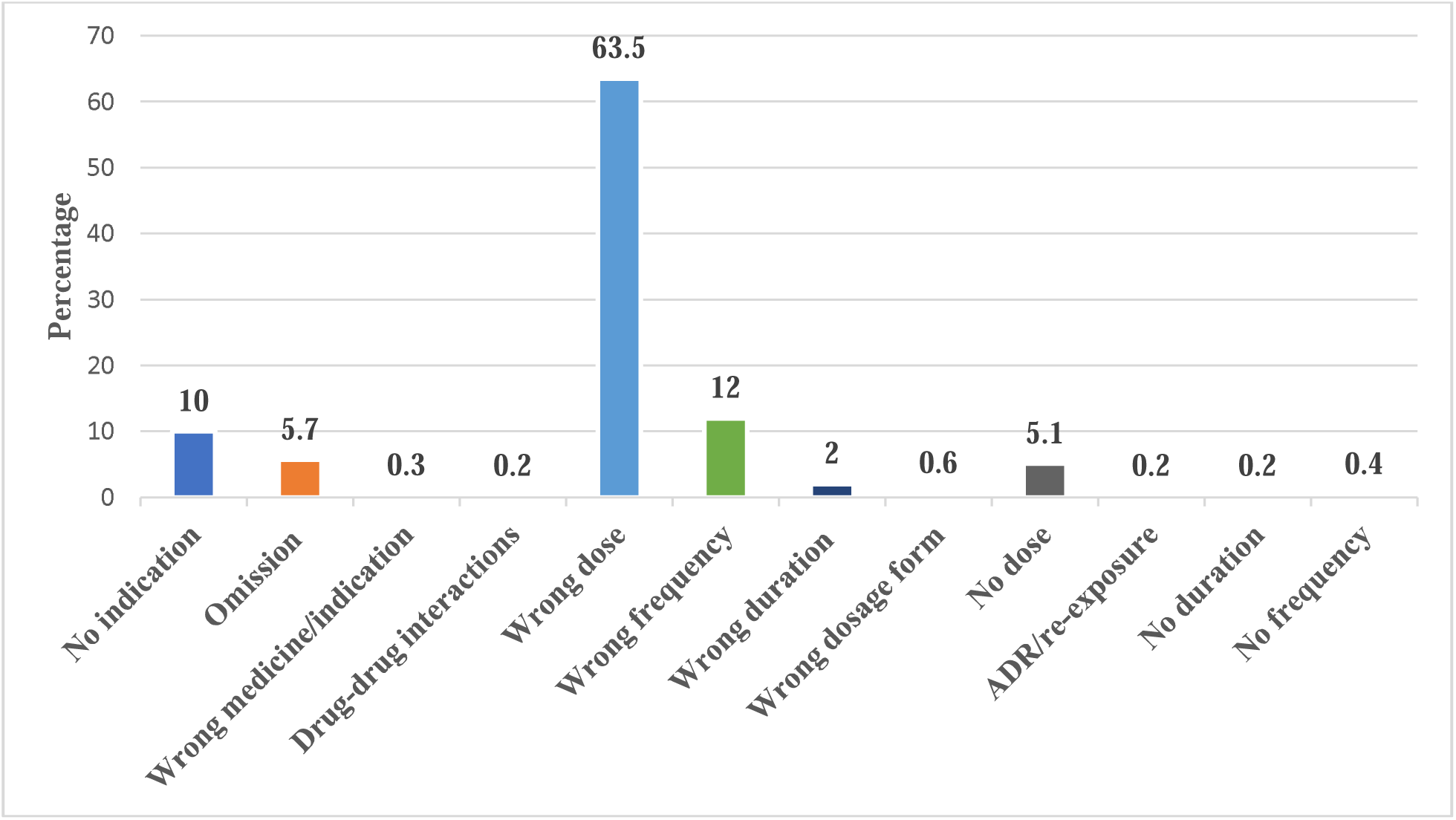
Proportion of different types of prescribing error at the outpatient setting, JDWNRH.

**Table 1.**
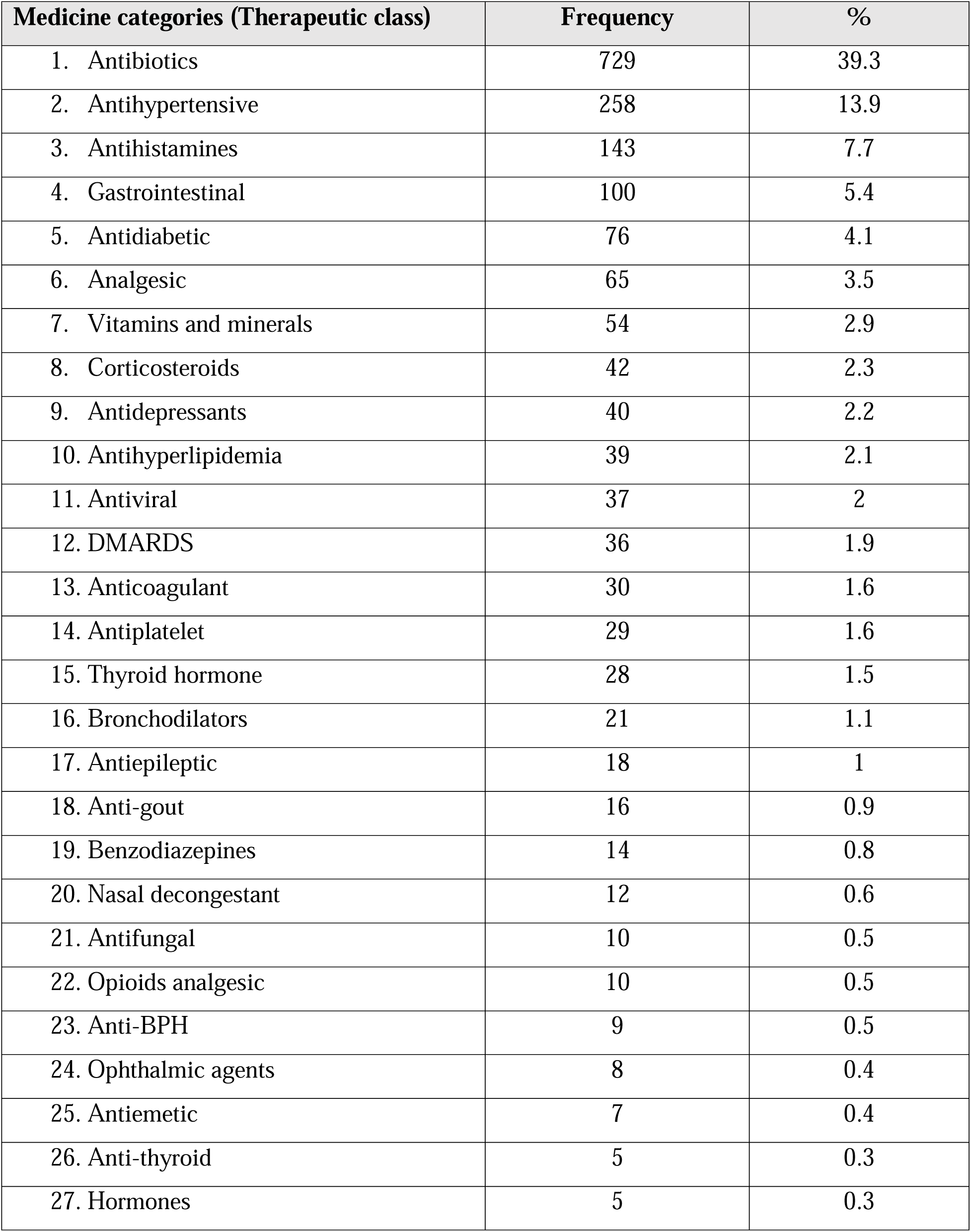
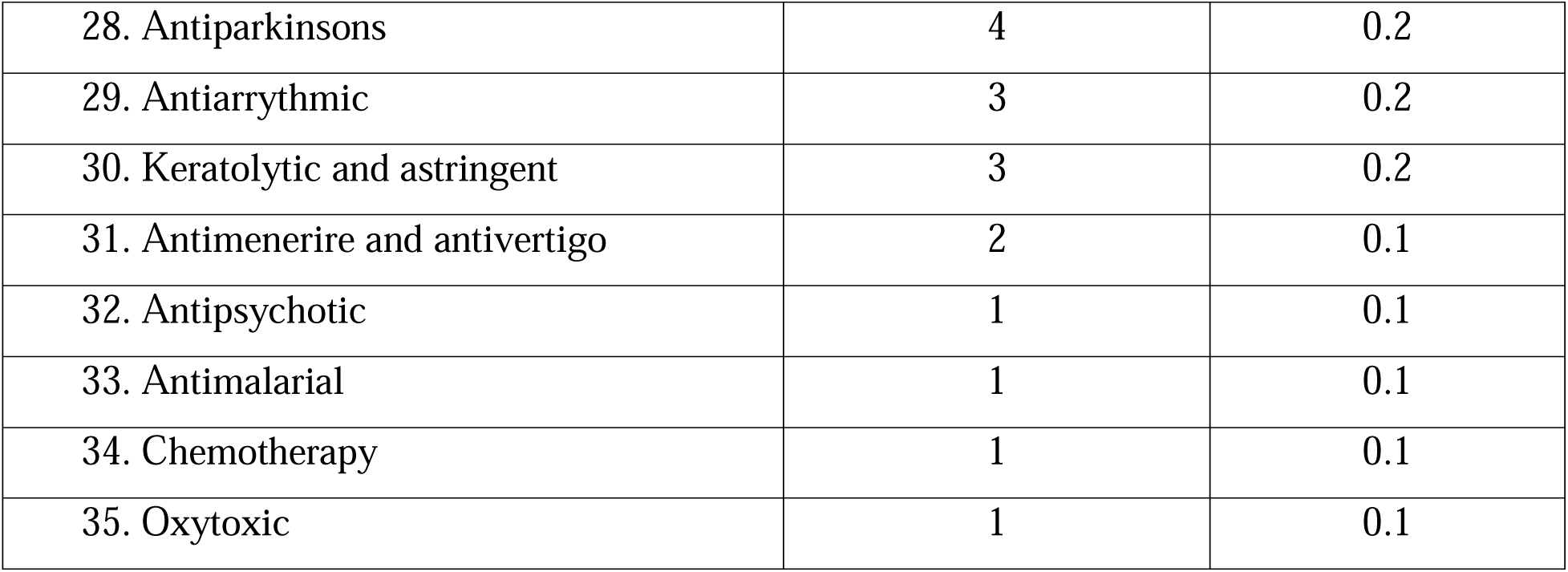
Ranking in therapeutic categories of medicines attributed with prescribing error at the outpatient setting, JDWNRH.

As shown in the **Figure 3**, the specialist (52%) and General Duty Medical Officer (GDMO) (27.1%) were the common prescribers who contributed to the overall prescribing error. In terms of department distribution, more than half were represented by the general outpatient (28.7%) and medical department (24.2%) combined as reflected in **Figure 4**.

**Figure 3.**
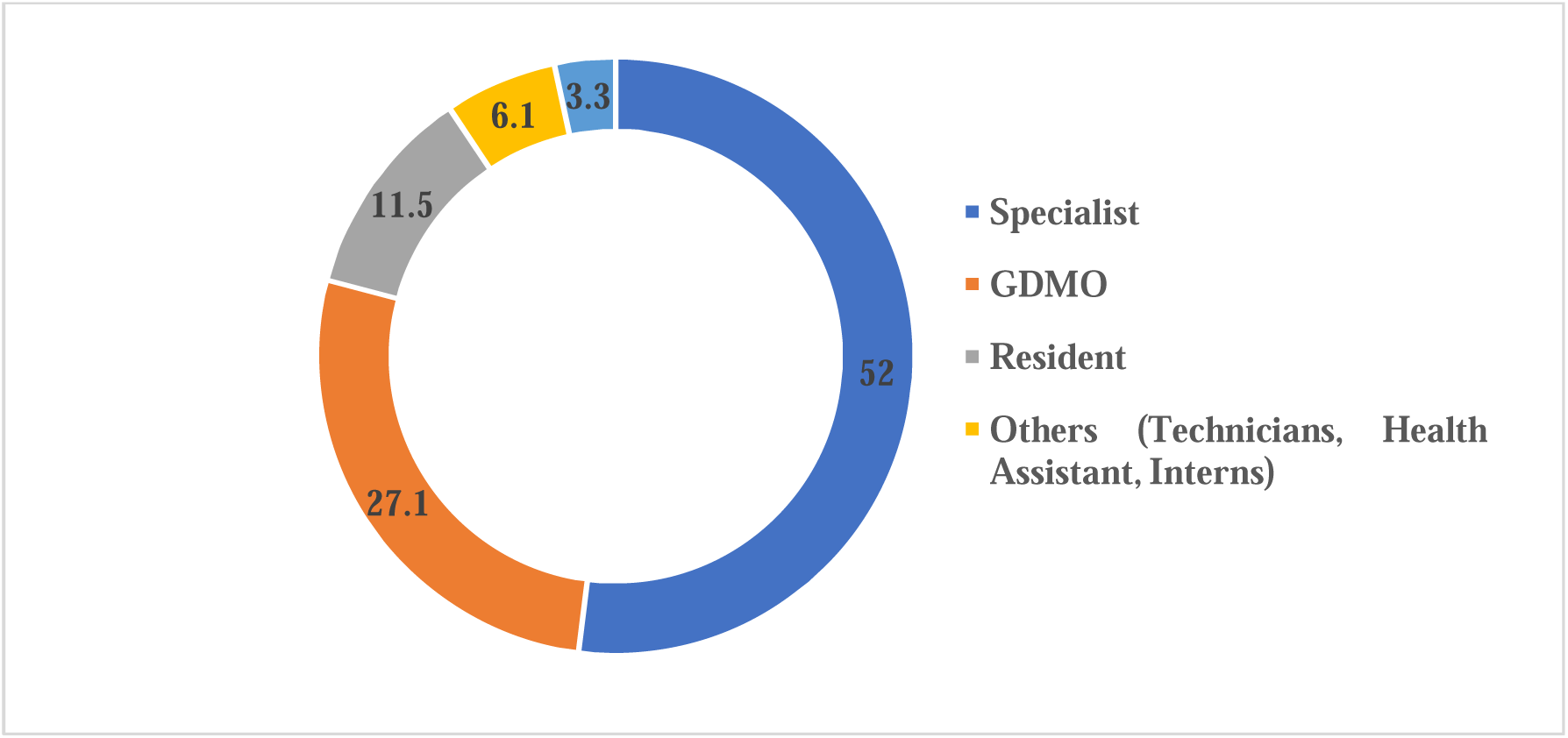
Proportion of prescriber categories who contributed to prescribing error at the outpatient setting, JDWNRH.

**Figure 4.**
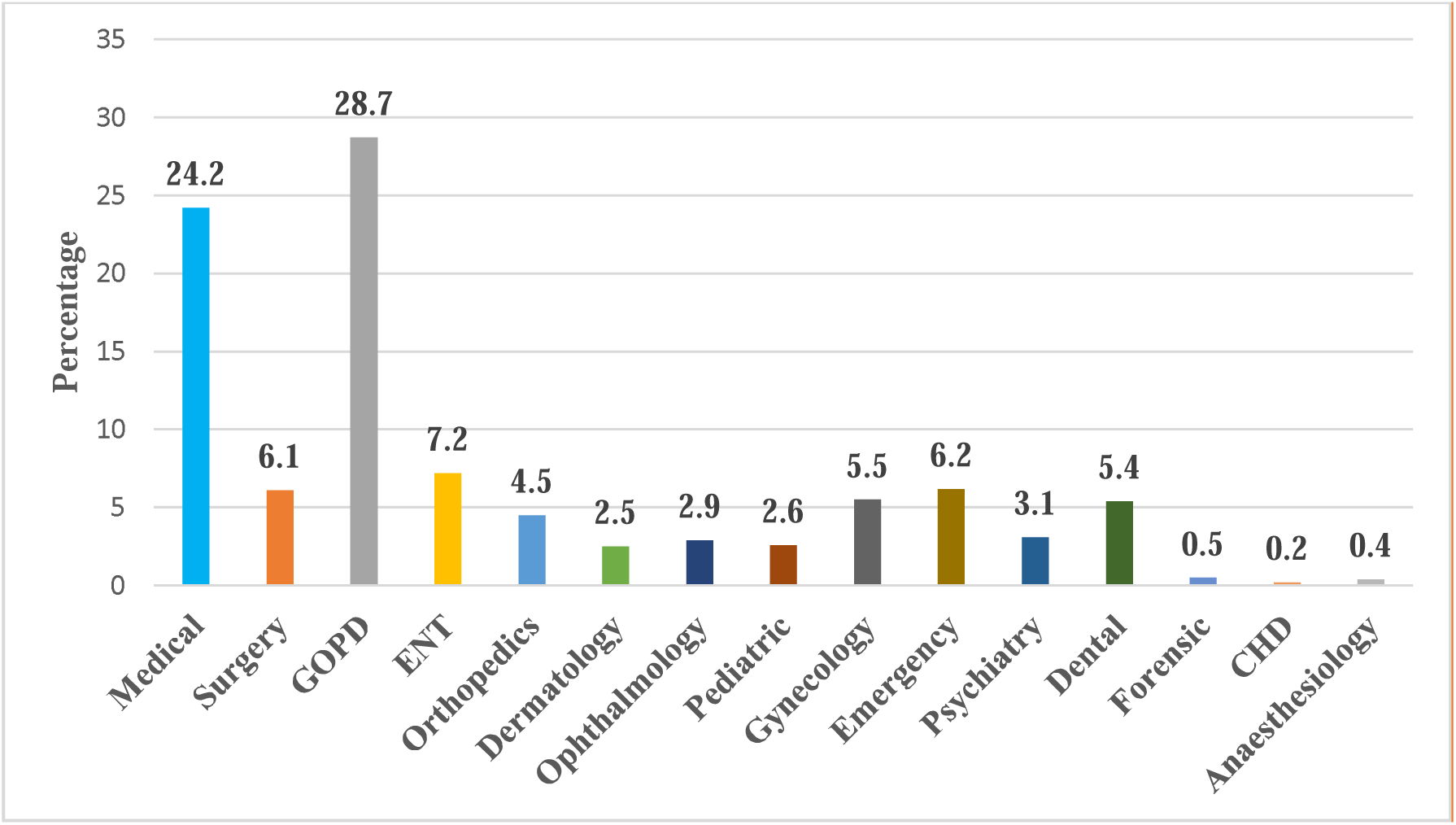
Proportion of departments who contributed to prescribing error at the outpatient setting, JDWNRH.

### Packaging and dispensing error

Of the 178 packaging errors, the common packaging error detected were antihypertensives and antidepressants with 30 (16.85%) and 26 (14.61%), respectively. All packaging errors were prevented by the dispensers from reaching it to the patients and no actual harm was associated with the packaging error being reported in our study. However, 100% of the dispensing error reported has reached to patients with antihypertensives being the most common category of medicine as described in **Figure 5**.

**Figure 5.**
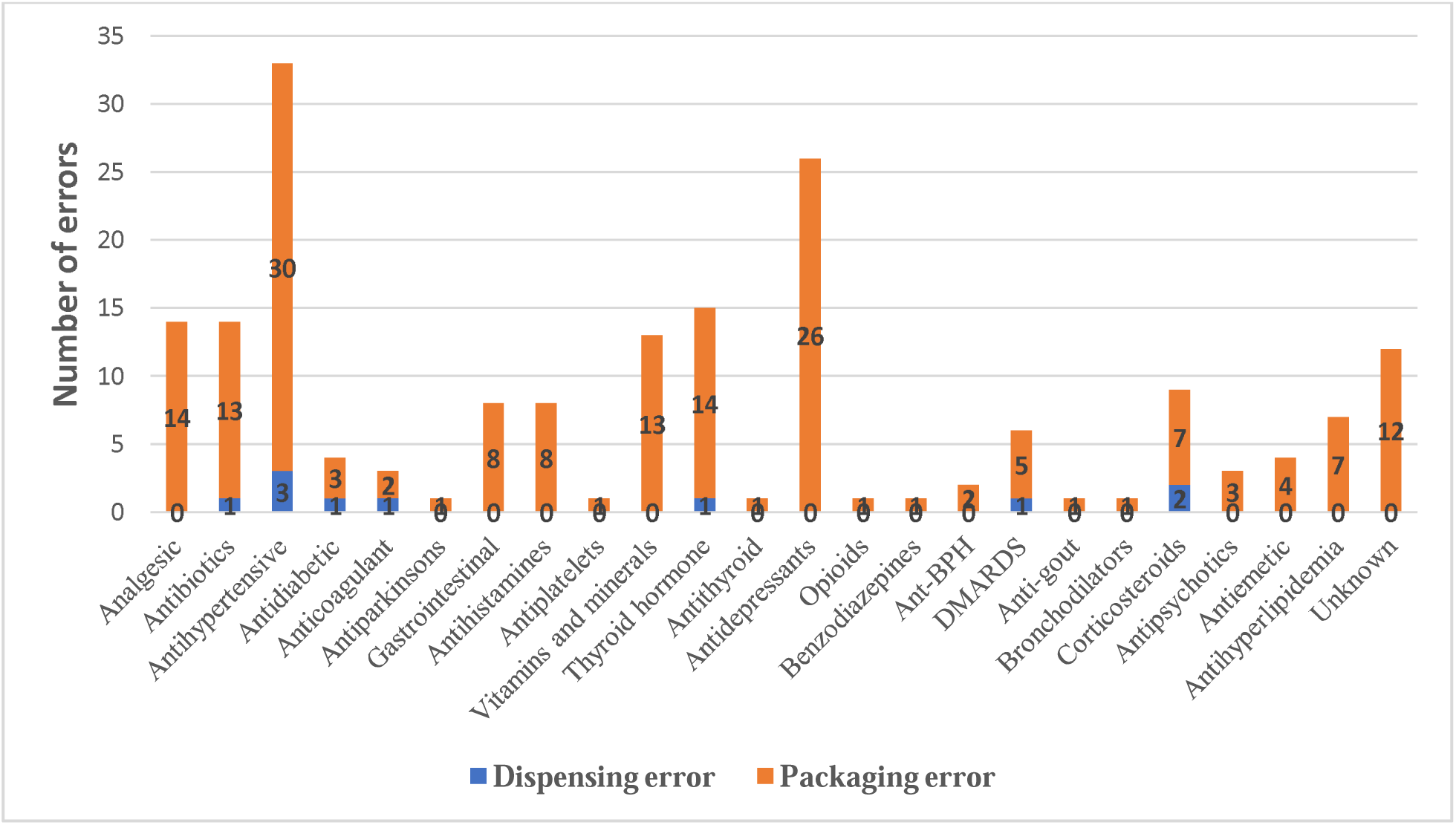
Number of packaging and dispensing errors at the outpatient setting, JDWNRH.

### Severity of harm associated with prescribing error

The severity of harm and the therapeutic classes of medicines are described according to the HAMEC tool as shown in **Table 2**. The most common severity of potential harm associated with the prescribing error was found to be moderate (1364, 73.45%), followed by minor (233, 12.55%), no harm (212, 11.42%) and serious (48, 2.58%). As depicted in **Table 3**, the actual harm that reached to patients were 8.96%, 6.87%, 6.23% and 8.33% for no harm, minor, moderate and serious, respectively.

**Table 2.** Potential harm associated with the prescribing error at the outpatient setting, JDWNRH.

| Severity of harm | Category of medicines | n (%) | Reached to patients? |  |
| --- | --- | --- | --- | --- |
|  |  |  | No | Yes |
| **No harm** | Antihistamines | 143 | 131 (91.61) | 12 (8.39) |
|  | Vitamins and minerals | 54 | 48 (88.89) | 6 (11.11) |
|  | Keratolytic and astringent | 3 | 3 (100.00) | 0 (0.00) |
|  | Nasal decongestant | 12 | 11 (91.67) | 1 (8.33) |
| **Minor** | Analgesic | 65 | 61 (93.85) | 4 (6.15) |
|  | Gastrointestinal | 100 | 94 (94.00) | 6 (6.00) |
|  | Ophthalmic | 8 | 6 (75.00) | 2 (25.00) |
|  | Antimenerire and antivertigo | 2 | 2 (100.00) | 0 (0.00) |
|  | Antiemetic | 7 | 7 (100.00) | 0 (0.00) |
|  | Corticosteroids | 42 | 39 (92.86) | 3 (7.14) |
|  | Anti-BPH | 9 | 8 (88.89) | 1 (11.11) |
| **Moderate** | Bronchodilators | 21 | 21 (100.00) | 0 (0.00) |
|  | Antipsychotic | 1 | 1 (100.00) | 0 (0.00) |
|  | Antimalarial | 1 | 1 (100.00) | 0 (0.00) |
|  | Hormones | 5 | 5 (100.00) | 0 (0.00) |
|  | Chemotherapy | 1 | 1 (100.00) | 0 (0.00) |
|  | Oxytoxic | 1 | 1 (100.00) | 0 (0.00) |
|  | Antihyperlipidemia | 39 | 36 (92.31) | 3 (7.69) |
|  | DMARDS | 36 | 33 (91.67) | 3 (8.33) |
|  | Anti-gout | 16 | 16 (100.00) | 0 (0.00) |
|  | Antibiotics | 729 | 687 (94.24) | 42 (5.76) |
|  | Antifungal | 10 | 9 (90.00) | 1 (10.00) |
|  | Antiviral | 37 | 37 (100.00) | 0 (0.00) |
|  | Antihypertensive | 258 | 237 (91.86) | 21 (8.14) |
|  | Antidiabetic | 76 | 71 (93.86) | 5 (6.58) |
|  | Antidepressants | 40 | 37 (92.50) | 3 (7.50) |
|  | Thyroid hormone | 28 | 25 (89.29) | 3 (10.71) |
|  | Anti-thyroid | 5 | 5 (100.00) | 0 (0.00) |
|  | Opioids analgesic | 10 | 9 (90.00) | 1 (10.00) |
|  | Benzodiazepines | 14 | 13 (92.86) | 1 (7.14) |
|  | Antiarrhythmic | 3 | 3 (100.00) | 0 (0.00) |
|  | Antiparkinsons | 4 | 4 (100.00) | 0 (0.00) |
|  | Antiplatelet | 29 | 27 (93.10) | 2 (6.90) |
| **Serious** | Anticoagulant | 30 | 27 (90.00) | 3 (10.00) |
|  | Antiepileptic | 18 | 17 (94.44) | 1 (5.56) |

**Table 3.**
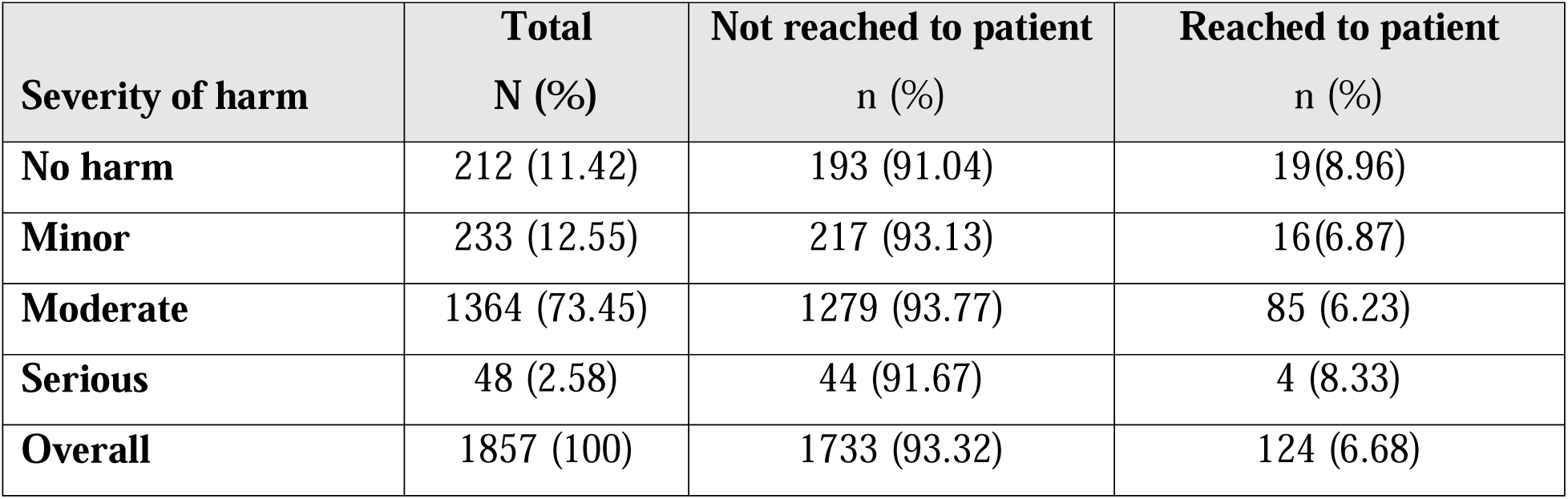
The overall proportion of the severity of potential and actual harm associated with prescribing error at the outpatient setting, JDWNRH.

| **Severity of harm** | **Total**<br>**N (%)** | **Not reached to patient**<br>**n (%)** | **Reached to patient**<br>**n (%)** |
| --- | --- | --- | --- |
| **No harm** | 212 (11.42) | 193 (91.04) | 19(8.96) |
| **Minor** | 233 (12.55) | 217 (93.13) | 16(6.87) |
| **Moderate** | 1364 (73.45) | 1279 (93.77) | 85 (6.23) |
| **Serious** | 48 (2.58) | 44 (91.67) | 4 (8.33) |
| **Overall** | 1857 (100) | 1733 (93.32) | 124 (6.68) |

## Discussion

### Prevalence of MEs

The findings from this study represents the prevalence and characteristics of the MEs at the outpatient care at JDWNRH, Bhutan. Considering the number of prescriptions dispensed as a reporting unit, the overall prevalence of MEs at the outpatient care at JDWNRH account to 2.27%.

Depending on the reporting system, a wide variation in the prevalence of overall MEs are being documented. For example, high prevalence of MEs ranging between 42% and 56% were reported [13,14]. Moreover, the incidence of MEs was reported as 12.5 per persons-year (95% CI 9.4-16.2) [15]. Therefore, the overall prevalence of MEs reported is primarily dependent on the definition of MEs used in the study.

Among different categories of MEs assessed, prescribing error (2.06%) was the most common type of error, followed by packaging error (0.20%) and dispensing error (0.01%). A similar study found that 7.8% of the prescriptions have prescribing errors among 2000 patients attending general practice clinic [16].

From the overall MEs, prescribing error, packaging error and dispensing error accounts to 90.81%, 8.70% and 0.49%, respectively. This is concurrent with the findings from a recent systematic analysis, where prescribing error at the outpatient and ambulatory settings was reported up to 91% among overall MEs assessed [17]. However, studies have also reported as the number of prescribing errors based on the categories of medicines with or without denominators [18,19]. This highlights the importance of considering the categories of medicines and method involved in reporting of MEs while making comparisons with other studies.

Among 24 studies included in the recent systematic review, none of the study reported the prevalence of packaging error [17]. Hence, the manual packaging of medicines using dispensing envelops by the pharmacy technicians represents a unique medication-use process involved in the outpatient care in Bhutan. Although the overall prevalence of packaging error was low (0.20%) and completely prevented from reaching it to the patients by the dispensers, our study doesn’t include packaging errors that has already reached the patients. Adopting a bar code reading and automated packaging system could prevent packaging error in our setting. Moreover, developing and implementing a standard operating procedure (SOP) on handling of look-alike medicines will minimize the overall packaging error in our study setting.

Dispensing error in our study was 0.01%. Our finding is lower than the study which focused on chemotherapy, with the dispensing error of 1.5% [20]. A study targeted in older patients detected the dispensing error incidence of 20.7/100 person-years [21].

The low prevalence of packaging and dispensing error in our study could be attributed to lack of active reporting by the patients. Such findings are supported by studies where lack and inefficient incident reporting system in healthcare organizations are attributed to under-reporting of MEs up to 60% [22,23]. Therefore, the overall prevalence of packaging and dispensing errors at outpatient care will be well represented if there is a standard incident reporting system accessible by patients to report errors when encountered.

### Characteristics of prescribing error

Based on the therapeutic classification, the most common medications attributed with the prescribing error was antibiotics (39.3%) and antihypertensive agents (13.9%). Similar findings were reported, where antibiotics (16.4%) [24] and antihypertensives (12.0%) [25], were the most common medicines involved in prescribing error. Antibiotics and antihypertensives being the most commonly prescribed medicines in the outpatient care could be linked to high prevalence of prescribing errors.

Dosing error (63.5%) was the most common type of prescribing error, followed by wrong frequency (12%), no indication (10.0%), omission (5.7%) and no dose (5.1%). Incorrect dosing is highly prevalent type of prescribing error reported from various healthcare facilities across the globe. In particular, a systematic review involving 24 studies documented dosing error ranging up to 41% of all medications prescribed [17]. A similar study reported 112 (25.5%) dosing errors in 479 prescriptions [26]. Lack of knowledge and inadequate training among prescribers related to recent therapeutic guidelines could be major contributory factors leading to overall prescribing errors. Similar conclusion was made by a recent systematic analysis which assessed factors associated with MEs [17]. However, errors such as no indication, omission and no dose mentioned could be attributed to lapses or failures of prescribers rather than their knowledge on therapeutic practices.

### Severity of prescribing error

According to HAMEC tool, our study concluded that the majority (73.45%) of the prescribing error was moderately severe. Among which, 93.77% and 6.23% were observed as potential and actual harm, respectively. Of the 1857 prescribing errors recorded, 1733 (93.32%) of the them were prevented from reaching it to the patients by the hospital pharmacist. This highlights the significant role of the hospital pharmacist in preventing the prescribing error in the outpatient setting. Furthermore, preventing majority of the prescribing error reflects the high acceptance level among concerned prescribers when intervened by the pharmacists. Accordingly, a pharmacist-led interventions has led to reduced prescribing errors and improved healthcare outcomes as reported by several studies [27,28]. On the other hand, about 8.33% (4/28) of the serious prescribing errors involving anticoagulant and antiepileptic have reached to patients, inducing serious harm in our study. However, our study failed to describe the actual harm associated with the serious prescribing error. Hence, there is a need to adopt a standard system where patients can be followed up and adverse drug effects associated with the prescribing error were managed appropriately.

## Conclusion

Our study represents the baseline data on the prevalence and characteristic of MEs at the outpatient setting in JDWNRH. Despite various definition of MEs used across the studies, the overall prevalence of MEs was low in our study setting. Prescribing error was the most common type of MEs recorded, where dosing error was highly prevalent type of prescribing error documented. Using the therapeutic classifications, more than half of the prescribing error was related to antibiotics and antihypertensives combined. Appropriate training on recent therapeutic guidelines, particularly infectious disease and hypertension management guidelines are highly recommended to minimize the overall prescribing errors.

After assessing the severity of prescribing error, majority of the errors were found to be moderate. On the positive note, significant proportion of prescribing errors were prevented from inducing actual harm among patients by the hospital pharmacists. This clearly highlights the importance of pharmacist-led interventions in preventing prescribing errors and its associated harm at the outpatient settings. In general, a multidisciplinary approach with effective integration of hospital pharmacist in disease management at outpatient setting will significantly enhance the overall patient safety.

Not being able to capture the demographic and clinical characteristics of the patients to assess the factors contributing to prescribing errors was the major limitation of our study. Lack of detailed description of MEs is another limitation of the study.

## Data Availability

All data produced in the present study are available upon reasonable request to the authors

## Acknowledgement

The authors would like to thank all the PT working at the department of pharmacy, JDWNRH for their contribution in the study.

